# Inactivation of highly pathogenic avian influenza virus with high temperature short time continuous flow pasteurization and virus detection in bulk milk tanks

**DOI:** 10.1101/2024.07.01.24309766

**Authors:** Erica Spackman, Nathan Anderson, Stephen Walker, David L. Suarez, Deana R. Jones, Amber McCoig, Tristan Colonius, Timothy Roddy, Nicholas J. Chaplinski

**Affiliations:** Exotic and Emerging Avian Viral Disease Research Unit, Southeast Poultry Research Center, US National Poultry Research Center, USDA-Agricultural Research Service, Athens, GA, USA; Office of Food Safety, Center for Food Safety and Applied Nutrition, US Food and Drug Administration, Bedford Park, IL, USA; US National Poultry Research Center, USDA-Agricultural Research Service, Athens, GA, USA; Center for Veterinary Medicine, US Food and Drug Administration, Rockville, MD, USA; Office of State Cooperative Programs, US Food and Drug Administration, Kansas City, MO, USA

**Keywords:** Influenza A, highly pathogenic avian influenza, raw milk, cows, pasteurization, high temperature short time (HTST)

## Abstract

Infections of dairy cattle with clade 2.3.4.4b H5N1 highly pathogenic avian influenza virus (HPAIV) were reported in March 2024 in the U.S. and viable virus was detected at high levels in raw milk from infected cows. This study aimed to determine the potential quantities of infectious HPAIV in raw milk in affected states where herds were confirmed positive by USDA for HPAIV (and therefore were not representative of the entire population), and to confirm that the commonly used continuous flow pasteurization using the FDA approved 72°C (161°F) for 15 s conditions for high temperature short time (HTST) processing, will inactivate the virus. Double-blinded raw milk samples from bulk storage tanks from farms (n=275) were collected in four affected states. Samples were screened for influenza A using quantitative real-time RT-PCR (qrRT-PCR) of which 158 (57.5%) were positive and were subsequently quantified in embryonating chicken eggs. Thirty-nine qrRT-PCR positive samples (24.8%) were positive for infectious virus with a mean titer of 3.5 log_10_ 50% egg infectious doses (EID_50_) per mL. To closely simulate commercial milk pasteurization processing systems, a pilot-scale continuous flow pasteurizer was used to evaluate HPAIV inactivation in artificially contaminated raw milk using the most common legal conditions in the US: 72°C (161°F) for 15s. Among all replicates at two flow rates (n=5 at 0.5L/min; n=4 at 1L/min), no viable virus was detected. A mean reduction of ≥5.8 ± 0.2 log_10_ EID_50_/mL occurred during the heating phase where the milk is brought to 72.5°C before the holding tube. Estimates from heat-transfer analysis support that standard U.S. continuous flow HTST pasteurization parameters will inactivate >12 log_10_ EID_50_/mL of HPAIV, which is ~9 log_10_ EID_50_/mL greater than the mean quantity of infectious virus detected in raw milk from bulk storage tank samples. These findings demonstrate that the milk supply is safe.

## Introduction

Highly pathogenic avian influenza virus (HPAIV) was reported in dairy cow herds in the U.S. in March of 2024 and virus was subsequently detected in raw milk *(Burrough et al., 2024)*. An April 2024 targeted study of retail fluid milk and dairy products reported that virus could be detected by quantitative real-time reverse transcription polymerase chain reaction (qrRT-PCR) based methods in approximately 20% of the products but no infectious virus was detected *(Spackman et al., 2024)*. Therefore, it is critical to understand the prevalence and potential quantities of infectious virus that could occur along each stage of the milk supply.

Bulk tanks are large storage tanks used to cool and store milk from the herd on a dairy farm until it can be picked up for processing. Larger farms can pump directly to tanker trucks to facilitate transfer to the processing plant. Tanks and tanker trucks have the capacity to hold milk from 600-700 cows. After transport to processing plants, milk is clarified and filtered, then separated so the milk fat can be standardized to the required content for the final product. Finally, the standardized product is homogenized at high pressure, which requires heating to 55-80°C. Homogenization occurs in-line with pasteurization, so the two processes occur together continuously.

Cows on larger farms are also closely monitored for feed consumption, milk production, and rumen activity; sick cows are separated for further attention. Cows with clinical mastitis, cows with abnormal milk quality, or sick cows undergoing treatment are milked separately so that the milk can be diverted from entering the human food supply. However, individual HPAIV infected cows that appear healthy have been observed *(Ashby, 2024)* and could potentially excrete virus into their milk, and cows that recover from infection and are returned to the general population may still shed viral RNA for extended periods of time.

Pasteurization, which uses heat to kill pathogenic bacteria, has been a public health success to greatly reduce human infections related to the consumption of dairy products *(FDA, 2019)*. Previously, there have been no studies on inactivation of avian influenza virus in milk, but based on pasteurization times in egg product it was expected that HPAIV in milk would be inactivated at common pasteurization times and temperatures *(Chmielewski et al., 2011;Chmielewski et al., 2013;Thomas and Swayne, 2009)*. However, recent studies attempting to simulate pasteurization conditions at the benchtop scale have reported inconsistent results. Guan *et al*., described a 4.5 log_10_ 50% tissue culture infectious dose reduction after 15 s at 72°C but infectious virus could be recovered beyond 15 s from milk with higher initial titers *(Guan et al., 2024)*. Similarly, another study reported a reduction of 4 log_10_ TCID_50_/mL in 5 s at 72°C, and infectious virus could still be detected to 20 seconds *(Kaiser et al., 2024)*. In contrast to the other studies, another found complete inactivation at 72°C in 15 s with log reductions up to 7.75 log_10_ EID_50_ /mL of an H5 virus in raw milk was reported *(Cui et al., 2024)*. As the authors of these reports recognize, because these studies were conducted using PCR thermocyclers, the studies do not directly replicate commercial pasteurization conditions. Therefore, the objectives of this study were to determine the potential quantities of infectious HPAIV in bulk tank milk and to assess the efficacy of continuous flow pasteurization under conditions that closely approximate commercial milk pasteurization processing at high temperature short time (HTST), 72°C (161°F) for 15 s.

## Materials and Methods

### Collection of bulk tank milk samples

In collaboration with four states, the U.S. Food and Drug Administration (FDA) secured raw milk bulk tank samples over a two-week period April 18-27, 2024. Samples were taken from farms known to be affected as well as farms not known to be affected. Samples were collected as “universal samples” as a routine procedure anytime milk was offered for sale, by licensed personnel, as part of state participation in the Pasteurized Milk Ordinance *(FDA, 2019)* regulatory system. Each sample was blinded and sent by state regulatory personnel to the Institute for Food Safety and Health (IFSH, Bedford Park, IL) where they were double-blinded and aggregated prior to shipment to the U.S. National Poultry Research Center (USNPRC) for analysis. These states had herds with HPAIV H5N1 infections confirmed by USDA. Sampling occurred in regions known to include affected farms and was neither random nor representative of prevalence.

### Quantitative real-time RT-PCR for influenza A

Upon receipt, double-blinded bulk tank samples were assigned a unique accession number and processed for RNA extraction as described using a hybrid procedure with Trizol LS (Thermo Fisher Scientific, Waltham, MA) and the MagMax magnetic bead kit *(Spackman et al., 2024)*. VetMAX Xeno (Thermo Fisher Scientific) was added to the Trizol LS for each reaction prior to sample addition to serve as an extraction control and an internal positive control. Each sample was run on quantitative real-time RT-PCR (qrRT-PCR) assay on a QuantStudio5 (Thermo Fisher Scientific) using a test targeting the influenza A M gene *(Spackman et al., 2002)*. The primers and probe for the internal control were used as directed by the Xeno kit instructions. Non-infectious qrRT-PCR based quantity estimates were determined by including a standard curve derived from RNA extracted from a 10-fold dilutions series of quantified avian influenza virus stocks propagated in embryonating chicken eggs *(Spackman, 2020)*.

### Virus detection and quantification in embryonating chickens eggs

All samples that were positive for virus detection by qrRT-PCR were processed to determine the quantity of infectious virus in embryonating chicken eggs (ECE). A portion of each sample (1 mL) was aliquoted and treated for 1 h at ambient temperature (approximately 21°C) with antibiotics at a final concentration of: penicillin G 1000 IU/ml, streptomycin 200 µg/ml, gentamicin100 µg/ml, kanamycin 65 µg/ml, amphotericin B 2 µg/ml. To quantify the virus, 10-fold dilutions were made in brain heart infusion (BHI) broth with antibiotics using standard methods *(Reed and Muench, 1938;Spackman and Killian, 2020)* and hemagglutination assay was used to confirm the presence of avian influenza virus *(Killian, 2020)*.

### Inactivation of HPAIV by HTST continuous flow pasteurization

Raw milk (approximately 4.5% milk fat) (Supplementary Table 1) was obtained from the University of Georgia Dairy (Athens, GA) and immediately transported to the USNPRC for processing. Milk was homogenized in a Gaulin 15m 8BA (Manton-Gaulin Manufacturing CO., Inc, Everett, MA) at 40°C. After homogenization, a 3mL sample was collected as a negative control and 5L portions of milk were prepared and artificially contaminated with a recent US clade 2.3.4.4b HPAIV isolate: A/turkey/Indiana/22-003707-003/2022 H5N1 (TK/IN/22) (provided by Dr. Mia Torchetti, National Veterinary Services Laboratories, US Department of Agriculture-Animal and Plant Health Inspection Service, Ames, IA).

A pilot-scale continuous flow pasteurizer modified for inline sampling and cooling (UHT/HTST Veros^TM^ EDH, MicroThermics®, Raleigh, NC) was installed in an ABSL-3Ag research space and used to closely simulate commercial milk pasteurization processing systems. The computer-controlled pasteurizer included a progressive cavity pump, preheater, final heater, hold tube, cooler, and inline sampling and cooling ports after the final heater and after the product cooler (Figure 1). Temperature was measured for critical parameters using calibrated thermocouples and recorded in 10 s intervals by the MicroThermics® system.

**Figure 1.**
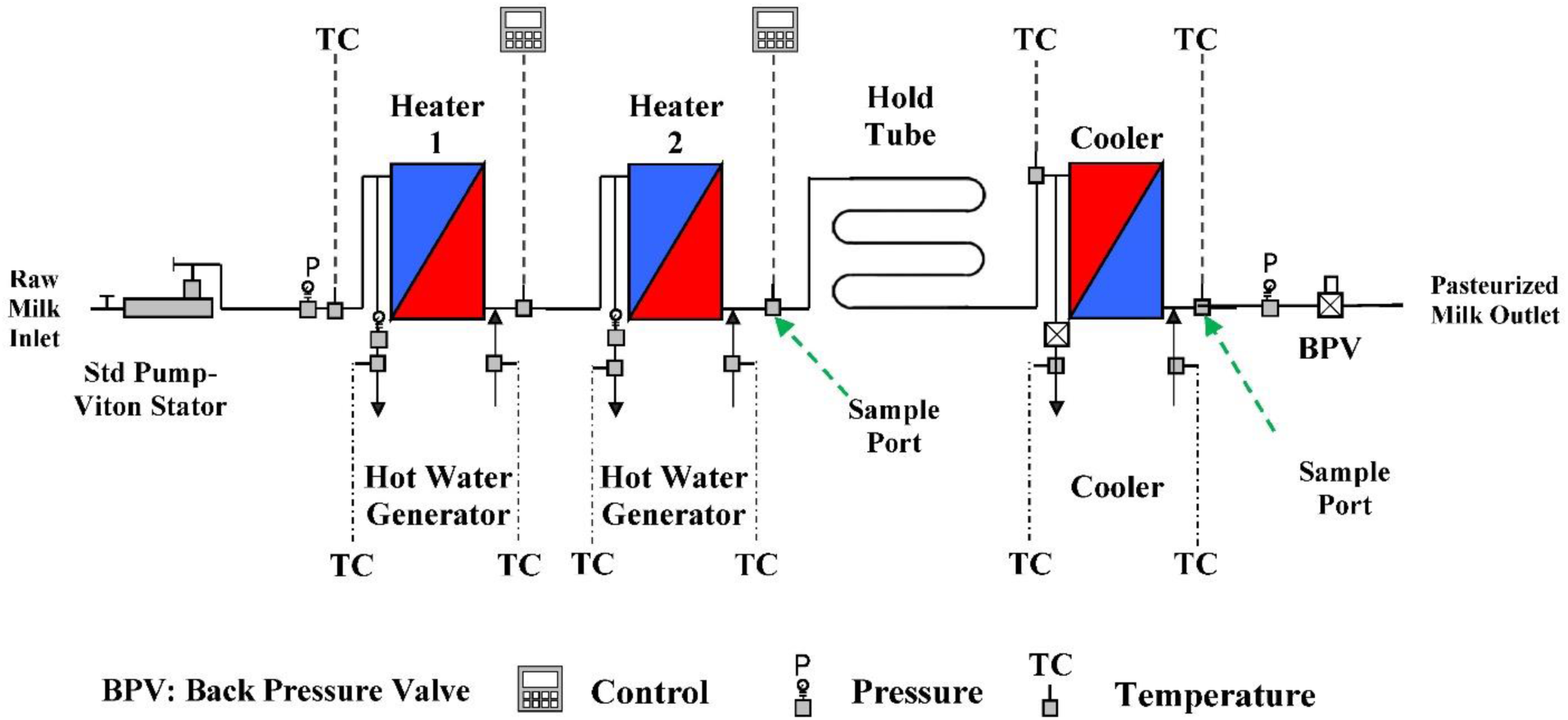
Schematic of the HTST continuous flow pasteurizer with inline sampling ports.

The artificially contaminated raw, homogenized milk (10 mL of virus stock added to 5 L raw milk at a titer of ~9.7 log_10_ EID_50_/mL) was used to supply the pasteurizer at a flow rate of 0.5 or 1.0 L/min. Flow rate was verified prior to each pasteurization run using a stopwatch and graduated cylinder. Milk was heated to 37.8°C in the preheater to ensure milk entered the final heater at a consistent temperature. Milk exited the final heater at 72.5°C and entered the hold tube, the section of the processor where the product is held for a specific time at a minimum temperature to achieve pasteurization. At a flowrate of 0.5 L/min, the product in the hold tube had a 15 s fastest particle residence time (FPRT) and 18 s bulk mean residence time (BMRT) and exited the hold tube at approximately 72°C, which constitutes the commonly used legal pasteurization treatment known as HTST (at least 72°C for minimum of 15 s). The higher flow rate of 1 L/min produced a sub-legal thermal treatment with a FPRT of only 7.5 s. Milk was cooled to approximately 21 to 32°C in the product cooler and excess milk was collected in sealed buckets for treatment with disinfectant.

Each sample set was collected on a different day starting with a fresh batch of raw milk from the dairy. Prior to each pasteurization run, three independent 3-mL samples were collected from the milk supplying the pasteurizer to establish the starting titer. During pasteurization run, two or three samples were collected at the outlet of the final heater, just before the product entered the hold tube. The inline sampling port cooled the milk in an ice-water bath prior to dispensing into a pre-sterilized septum bottle. Two or three samples were collected after the hold-tube at the outlet of the cooler (Table 1). All samples were quantified in ECE as described above. At least four replicate trials were completed at each flow rate.

**Table 1.** Summary of HTST continuous flow pasteurization parameters and quantities of infectious highly pathogenic avian influenza virus (HPAIV) in raw homogenized milk supplying the pasteurizer determined by a viability assay conducted in embryonating chicken eggs. Five and four replicates were conducted at flow rates of 0.5 L/min and 1 L/min, respectively. Values reported as mean ± standard deviation.

| **Milk**<br>**Flowrate**<br>**(L/min)** | **Raw Milk**<br>**Temperature**<br>**°C (°F)** | **Preheater**<br>**Temperature**<br>**°C (°F)** | **Final Heater**<br>**Temperature**<br>**°C (°F)** | **Hold Tube**<br>**Outlet**<br>**Temperature**<br>**°C (°F)** | **Titer of HPAIV**<br>**in supply milk**<br>**(Log<sub>10</sub>**<br>**EID<sub>50</sub>/mL)<sup>a</sup>** |
| --- | --- | --- | --- | --- | --- |
| 0.5 | 30.11 ± 0.17 | 39.84 ± 0.18 | 72.54 ± 0.16 | 71.64 ± 0.27 | 6.6 ± 0.15 |
| | (86.20 ± 0.32) | (103.71 ± 0.31) | (162.58 ± 0.30) | (160.96 ± 0.50) | |
| 1 | 31.43 ± 0.26 | 40.29 ± 0.33 | 72.71 ± 0.15 | 72.26 ± 0.16 | 6.8 ± 0.17 |
| | (88.57 ± 0.45) | (104.52 ± 0.64) | (162.87 ± 0.27) | (162.07 ± 0.28) | |

a. Mean of all replicates for each flow rate; EID<sub>50</sub> = 50% Egg infectious doses

Alkaline phosphatase activity measured soon after pasteurization is routinely used as an indicator to confirm that proper minimum pasteurization times and temperatures have been achieved (Rankin et al., 2010) and was used as an additional confirmation for differences in flow rate. An alkaline phosphatase test (Fast alkaline phosphatase test Charm Sciences, Inc, Lawrence, MA) read on the NovaLUM II-X (Charm Sciences, Inc.) was used in accordance with the manufacturer’s instructions on the supply milk, pre- and post-hold tube samples two replicates at 72°C 0.5 L/min and 1 L/min.

### Temperature Analysis of Final Heater

The final heater in the Microthermics® system is a coiled tube in shell heat exchanger consisting of 13.1 m of 6.35 mm x 0.89 mm (¼ in. x 0.035 in.) wall tubing. To help account for the lethality that occurs in the final heater section, it is useful to consider the temperature profile of the product as it passes through the heat exchanger. Using a few simple equations, the temperature profile across the length of the heater can be estimated. First, the heat transfer of a shell in tube heat exchanger is described by the following equation *(Heldman and Singh, 1981)*:

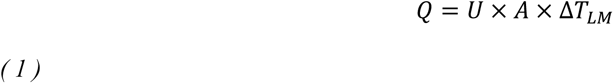

Where *Q* is the rate of heat transfer, *U* is the overall heat transfer coefficient, A is the heat transfer surface area, and Δ*T_LM_* is the log mean temperature difference between the product and the heating medium:

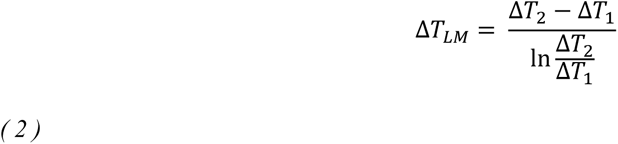

The rate of heat transfer in the heat exchanger is also equivalent to the rate of heat absorbed by the milk:

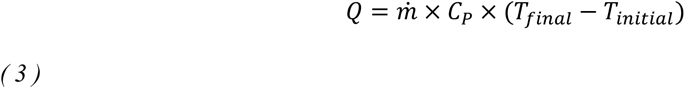

Where *ṁ* is the mass flow rate of milk, *C_P_* is the specific heat of milk, and (*T_final_* − *T_initial_*) is the temperature gained in the heat exchanger. Combining equation (1) and (3) and solving for *U*:

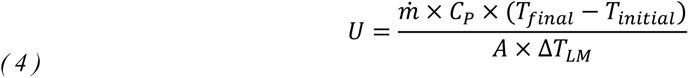

Using the reasonable assumption that the coefficient *U* is constant throughout the length of the heat exchanger, *U* can also be expressed in terms of any product temperature along the length of the heat exchanger:

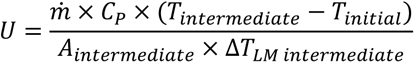

Combining equations (4) and (5) results in an expression that can be solved for the fraction of the heat exchanger area required to reach an intermediate temperature:

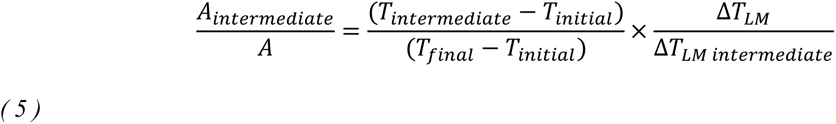

With a fixed cross-sectional area in the heat exchanger, this fraction also represents the fraction of the total residence time required to achieve an intermediate temperature. Because the temperature of the heating water is relatively constant across the heat exchanger, an estimate of residence time starting at any intermediate temperature can be calculated.

### Statistics

Simple linear regression was used to characterize the relationship between the qrRT-PCR estimated titer and actual infectious virus (Prism10.2, Graphpad Software, San Diego CA).

## Results

### Detection and quantification of influenza A in bulk tank milk samples

A total of 275 samples were tested for influenza A by qrRT-PCR and 158 (57.5%) were positive 107 (38.9%) were negative, and 10 (3.6%) were invalid (negative for influenza A and the internal control failed) (Supplementary Table 2). Of the 158 qrRT-PCR positive samples, one was discarded due to bacterial contamination and 39 (24.8%) were positive for infectious virus with titers from 1.3 to 6.3 log_10_ EID_50_/mL and a mean of 3.5 log_10_ EID_50_ /mL. There was no clear correlation between the estimated titer by qrRT-PCR and quantified viable virus (R-squared = 0.37) and the amount of live virus grown in ECE.

### Inactivation of HPAIV by continuous flow pasteurization

Processing conditions during pasteurization are reported in Table 1. Hold tube outlet temperature (the critical temperature parameter for pasteurization) was 71.64 ± 0.3°C (160.96 ± 0.50 °F) across all replicate trials at a flowrate of 0.5 L/min and 72.26 ± 0.15°C (162.07 ± 0.28°F) at 1.0 L per min. Input titers ranged from 6.4 to 7.1 log_10_ EID_50_/mL (mean 6.7 ± 0.2 log_10_ EID_50_/mL) (Table 1, Supplementary table 3). There were 5 replicates at 72°C, 0.5 L/min; four replicates at 72°C, 1 L/min.; and, 1 replicate each at 78, 83, 86, and 90°C at 0.5 L/min flow rate (Supplementary table 3). Regardless of target temperature and flow rate, no infectious virus was recovered from either sample location, before and after the hold tube. There consistently was an ≥ 5.8 log_10_ EID_50_ /mL reduction in titer of infectious virus at 72°C. Alkaline phosphatase was inactivated at 0.5 L/min, but failure to inactivate at the 1 L/min flow rate confirmed that the thermal treatment was reduced (Supplementary table 3).

## Discussion

When HPAIV was discovered in milk from infected cows *(Burrough et al., 2024)*, food safety concerns were raised as ingestion of contaminated milk could serve as a potential route of exposure for humans. Therefore, several points along the milk food supply chain were evaluated for the presence of infectious HPAIV using qrRT-PCR based methods and positive samples were then tested to quantify infectious virus. Retail milk products were shown to contain no infectious virus in a prior study *(Spackman et al., 2024)*, although 20% of the samples were positive for viral RNA. In the current study, two additional stages of the milk supply, bulk storage tanks and pasteurization, were assessed.

Because viral RNA has been detected in retail milk samples *(Spackman et al., 2024)*, its detection in bulk tanks was not unexpected. Interestingly, the proportion of samples that were positive for infectious virus was only 24.8% of the samples which were positive by qrRT-PCR. Also, the quantities of infectious virus were generally lower than what was detected by qrRT-PCR with a mean of 3.5 log_10_ EID_50_/mL. The discrepancy may be due to bacterial digestion of the virus and/or neutralizing antibodies from infected cows in the milk. Additional research on reliable assays for neutralizing influenza A antibody detection in raw milk are needed. Also, once cows recover from infection and return to normal milk production and quality, detectable, but non-infectious, viral RNA could be present in their milk (personal communication Dr. Mia Torchetti), but research is ongoing.

It is important to recognize that the first risk mitigation measure is to remove infected cows from contributing to the milk supply destined for commercial processing by PMO regulations. Based on field reports infected cows may develop mastitis or other clinical signs that will trigger diversion of the cow’s milk from the food supply. However, it is expected that early in infection, cows may be sub-clinical but have virus in the milk. Subclinical infections in cows may also play a role. There is also data to show that some cows may remain subclinical, but shed virus in the milk *(Ashby, 2024)*. Because identification of infected cows based on clinical signs cannot be 100% reliable, it is likely that virus contamination of milk will occur and other mitigation measures, such as pasteurization, are needed to assure a safe milk supply. Because the consumption of raw milk or dairy products is a known source of bacterial infections, the use of pasteurization has been widely adopted to kill pathogenic bacteria which greatly increases the safety of milk and dairy products. Because avian influenza has not been previously reported in milk, no safety data was available, but pasteurization methods have been reported in egg and egg products that support that temperatures similar to milk pasteurization are effective at inactivating avian influenza viruses *(Chmielewski et al., 2011;Chmielewski et al., 2013;Swayne and Beck, 2004;Thomas et al., 2008;Thomas and Swayne, 2009)*. However, recent studies appear to indicate that influenza A may have greater than expected thermal stability in milk and have reported somewhat inconsistent results for virus inactivation in raw milk at 72°C using PCR thermal cyclers *(Cui et al., 2024;Guan et al., 2024;Kaiser et al., 2024)*. Therefore, studies that more closely simulate commercial pasteurization are needed to ensure that continuous flow pasteurization, which is widely used by the U.S. dairy industry, is effective at eliminating infectious virus from milk.

An initial pasteurization run was conducted with 72, 78, 83, 86, 90°C at a 0.5 L /min flow rate and because no viable virus was recovered, all subsequent testing was conducted at the lowest temperature of 72°C. Importantly, no infectious virus was detected after the final heater (i.e., before the hold tube) where the milk was heated from 40 to 72.5°C (104 to 162.5°F). The inactivation of virus in the heating section makes it challenging to estimate the log reduction that might occur in the holding tube, although it would be expected to be much greater than the reduction achieved during the ramped heating process.

To estimate the lethality that would likely occur in the holding tube, consider that at 1.0 L per minute, the BMRT in the final heater is 12.9 s. Using the conservative estimates for turbulent flow, the FPRT in the center of tube is 9.9 s. At an intermediate temperature of 63°C (145°F), Equation 6 results in a fraction of 0.27, meaning that in the first 2.7 s the milk is heating from 40°C (104°F) to 63°C (145°F), and takes the remaining 7.2 s to reach 72.5°C (162.5°F). Data from a recently published study suggests that the decimal reduction time (D value) of HPAI in milk at 63°C (145°F) is approximately 20 s *(Kaiser et al., 2024)*, so it would be reasonable to assume that there is negligible reduction in infectious virus during the 2.7 s before it reaches 63°C (145°F), and therefore the entire inactivation of ~6 log_10_ EID_50_/mL is achieved during the 7.2 s of ramped heating from 63°C (145°F) to 72.5°C (162.5°F).

Commercial pasteurization systems in the U.S. have holding times that are calibrated to the FPRT using a salt-solution conductivity test *(FDA, 2019)*, so that the legal minimum holding times are achieved even for the fastest particle, as required under Title 21 of the Code of Federal Regulations (CFR) 1240.61 *(FDA, 1992)*. With inactivation of ≥5.8 log_10_ EID_50_ /mL in 7.2 seconds while heating to process temperature, it would be reasonable to expect that a 15 s holding time at a process temperature of 72°C (161°F) could inactivate >12 log_10_ EID_50_ /mL.

There are several limitations of these studies. First, the bulk tank sampling was limited in scope to milk from regions known to include affected farms and therefore was biased toward positive samples and was not designed to determine the prevalence of HPAIV in bulk tank milk at large (i.e., nationally representative sample). The sampling did provide a snapshot of the quantity of infectious virus that could be present in raw milk from affected farms. Second, challenge studies utilizing the continuous flow pasteurizer utilized high titers introduced into milk instead of milk from infected cows. High inoculum levels are typically used when conducting inactivation studies to document high levels of inactivation *(National Advisory Committee on Microbiological Criteria for, 2010)*. Naturally contaminated milk in quantities needed to complete replicates with the pasteurizer with consistent, high levels of HPAIV, was not available. Conducting challenge studies with inoculated product is common practice and results with naturally contaminated milk are expected to be similar, but studies with foot and mouth disease have shown that virus in milk from infected cows was more stable than virus in artificially contaminated milk *(Sellers, 1969)*. Because of the documented thermal lability of influenza A, and the de minimis thermal resistance exhibited by HPAIV in this study, any differences are expected to be minimal. Lastly, only whole milk fat content of approximately 4.5% was tested. Fat could protect the virus, so potentially virus could be more stable in higher-fat cream products *(Tomasula and Konstance, 2004)*. Because of the known protective effect of fat for bacterial pathogens, higher times and longer temperatures are required for higher fat products *(FDA, 2019;FDA, 2024)*. Additional studies are needed to accurately characterize the inactivation kinetics (D- and Z-value) of HPAIV in milk and milk products to assess process lethality under various time-temperature combinations.

This study demonstrated that infectious influenza A can be detected in bulk storage tanks from HPAIV infected dairy herds and that HTST pasteurization is effective in inactivating this virus in milk. Importantly, the quantities of infectious virus are generally much lower than what is detected by qrRT-PCR methods and no infectious virus could be detected in approximately 75% of the samples that were positive by qrRT-PCR. Approximately 5.8-6 log_10_ EID_50_ HPAIV was inactivated in the final heater before the holding tube (quantities of around 6-7 log_10_ viable units of an organism are typically used when conducting inactivation studies to document high levels of inactivation *(National Advisory Committee on Microbiological Criteria for, 2010)*). Also, the quantities of virus that were consistently inactivated in the final heater were approximately 3.0log_10_ EID_50_ higher than the mean quantity of infectious virus in this limited set of bulk tank samples. The pasteurization time-temperature combination in 21 CFR 1240.61 *(FDA, 1992)* and Pasteurized Milk Ordinance *(FDA, 2019)* of 72°C for 15s is estimated to result in >12 log reduction of HPAIV in whole milk under conditions that closely approximate HTST commercial milk pasteurization processing, which is further supported by the fact that retail milk products were shown to contain no infectious virus *(Spackman et al., 2024)*. These findings together demonstrate that the milk supply is safe. Additional work will evaluate the effect of homogenization on virus viability and the determination of D- and z-values for HPAIV in fluid dairy products.

## Data Availability

All data produced in the present study are available upon reasonable request to the authors

## Acknowledgements

The authors gratefully thank: Frankie Beacorn, Tarrah Bigler, Sydney Birge-Hales, Suzanne DeBlois, Edna Espinoza, Jesse Gallagher, Javier Garcia, Jessica Gladney, David Haley, Anne Hurley-Bacon, Lindsay Killmaster, Scott Lee, John Miles, David Miles, Stephen Norris, Phil Paxton, Jodie Ulaszek Melinda Vonkungthong, Robin Woodruff, Jason Wan, and Ricky Zoller for technical assistance with this work. We are also very grateful to the University of Georgia Dairy for supplying fresh raw milk, and The Pennsylvania State University, Department of Food Science for providing the homogenizer.

Any use of trade, firm, or product names is for descriptive purposes only and does not imply endorsement by the US Government. This research was supported by US Department of Agriculture (USDA)-Agricultural Research Service Project No. 6040-32000-081-00D and the US Food and Drug Administration by IAA 60-6040-4-005. All opinions expressed in this paper are the authors’ and do not necessarily reflect the policies and views of the USDA or FDA. The USDA and FDA are equal opportunity providers and employers.

## Abbreviations

BHI=brain heart infusion; BMRT = Bulk Mean Residence Time; CFR=Code of Federal Regulations; ECE = embryonating chicken egg; EID50= 50% egg infectious dose; FPRT = Fastest Particle Residence Time; HPAIV= highly pathogenic avian influenza virus; HTST = high temperature short time; IFSH = Institute for Food Safety and Health; qrRT-PCR=quantitative real-time reverse transcription polymerase chain reaction; 50% tissue culture infectious doses=TCID50. USNPRC=US National Poultry Research Center

## Supplementary materials

**Supplementary Table 1.** Analysis of milk used with the continuous flow pasteurizer. Raw milk was run on a LactiCheck™ Rapid Read Milk Analyzer (Weber Scientific, Hamilton, NJ), in accordance with the manufacturer’s instructions.

| Metric | Batch |  |  |
| --- | --- | --- | --- |
|  | 1 | 2 | 3 |
| Temperature (°C) | 17.5 | 16.0 | 18.6 |
| % Fat | 4.45 | 4.15 | 4.30 |
| % Solids not fat | 9.02 | 9.17 | 9.16 |
| Density (in 10XX.XX g/cm<sup>3</sup>) | 30.16 | 31.03 | 30.85 |
| Connectivity | 4.31 | 4.41 | 4.42 |
| % Water added | 0.00 | 0.00 | 0.00 |
| % Protein | 3.27 | 3.34 | 3.33 |
| Freezing point (°C) | -0.58 | -0.59 | -0.59 |
| % Solids, salt, others | 0.73 | 0.74 | 0.74 |
| % Total solids | 13.47 | 13.32 | 13.46 |
| % Lactose | 4.94 | 5.03 | 5.02 |
| pH | 6.5-7.0 | 6.5-7.0 | 6.5-7.0 |

**Supplementary table 3.** Summary of HPAIV inactivation in homogenized raw milk by continuous flow pasteurization.

| Batch<sup>a</sup> | Temp (°C) | Flow rate (L per min) | Titer of input milk<sup>b</sup> log<sub>10</sub> EID<sub>50</sub><sup>c</sup> | Replicate | Virus detection<sup>d</sup> log<sub>10</sub> EID<sub>50</sub> |  | Alkaline phosphatase test<sup>e</sup> |  |  |
| --- | --- | --- | --- | --- | --- | --- | --- | --- | --- |
|  |  |  |  |  | Pre-hold tube | Post-hold tube | Supply milk | Pre-hold tube | Post-hold tube |
| 1 | 72 | 0.5 | 6.8 | 1 | <0.8 | <0.9 | Not run | Not run | Not run |
|  |  |  |  | 2 | <0.9 | <0.9 | Not run | Not run | Not run |
|  |  |  |  | 3 | <0.9 | <0.9 | Not run | Not run | Not run |
| 1 | 78 | 0.5 | 6.9 | 1 | <0.9 | <0.9 | Not run | Not run | Not run |
|  |  |  |  | 2 | <0.9 | <0.9 | Not run | Not run | Not run |
|  |  |  |  | 3 | <0.9 | <0.9 | Not run | Not run | Not run |
| 1 | 83 | 0.5 | 6.6 | 1 | <0.9 | <0.9 | Not run | Not run | Not run |
|  |  |  |  | 2 | <0.9 | <0.9 | Not run | Not run | Not run |
|  |  |  |  | 3 | <0.9 | <0.9 | Not run | Not run | Not run |
| 1 | 86 | 0.5 | 6.8 | 1 | <0.9 | <0.9 | Not run | Not run | Not run |
|  |  |  |  | 2 | <0.9 | <0.9 | Not run | Not run | Not run |
|  |  |  |  | 3 | <0.9 | <0.9 | Not run | Not run | Not run |
| 1 | 90 | 0.5 | 7.1 | 1 | <0.9 | <0.9 | Not run | Not run | Not run |
|  |  |  |  | 2 | <0.9 | <0.9 | Not run | Not run | Not run |
|  |  |  |  | 3 | <0.9 | <0.9 | Not run | Not run | Not run |
| 2 | 72 | 0.5 | 6.5 | 1 | <0.9 | <0.9 | Not run | Not run | Not run |
|  |  |  |  | 2 | <0.9 | <0.9 | Not run | Not run | Not run |
| 2 | 72 | 0.5 | 6.6 | 1 | <0.9 | <0.9 | Not run | Not run | Not run |
|  |  |  |  | 2 | <0.9 | <0.9 | Not run | Not run | Not run |
| 3 | 72 | 0.5 | 6.6 | 1 | <0.9 | <0.9 | Fail | Pass | Pass |
|  |  |  |  | 2 | <0.9 | <0.9 | Not run | Fail | Pass |
| 3 | 72 | 0.5 | 6.4 | 1 | <0.9 | <0.9 | Fail | Pass | Pass |
|  |  |  |  | 2 | <0.9 | <0.9 | Not run | Fail | Pass |
| 2 | 72 | 1 | 6.5 | 1 | <0.9 | <0.9 | Not run | Not run | Not run |
|  |  |  |  | 2 | <0.9 | <0.9 | Not run | Not run | Not run |
| 2 | 72 | 1 | 6.8 | 1 | <0.9 | <0.9 | Not run | Not run | Not run |
|  |  |  |  | 2 | <0.9 | <0.9 | Not run | Not run | Not run |
| 3 | 72 | 1 | 6.9 | 1 | <0.9 | <0.9 | Fail | Fail | Fail |
|  |  |  |  | 2 | <0.9 | <0.9 | Not run | Fail | Fail |
| 3 | 72 | 1 | 6.8 | 1 | <0.9 | <0.9 | Fail | Fail | Fail |
|  |  |  |  | 2 | <0.9 | <0.9 | Not run | Fail | Fail |

a. Each batch is a different day run with fresh raw milk collected that morning.
b. Mean of 3 samples of input milk.
c. EID<sub>50</sub> = 50% Egg infectious doses.
d. Virus viability assay conducted in embryonating chicken eggs.
e. Test used to confirm that pasteurization times and temperatures have been achieved. Test kit: Fast alkaline phosphatase test (Charm Sciences, Inc, Lawrence, MA) read on the NovaLUM II-X (Charm Sciences, Inc.) in accordance with the manufacturer's instructions.

